# Immunological assays for SARS-CoV-2: an analysis of available commercial tests to measure antigen and antibodies

**DOI:** 10.1101/2020.04.10.20061150

**Authors:** John M. González, William J. Shelton, Manuel Díaz-Vallejo, Victoria E. Rodriguez-Castellanos, Juan Diego H. Zuluaga, Diego F. Chamorro, Daniel Arroyo-Ariza

## Abstract

The rapid spread of SARS-CoV-2 coronavirus infection has led to the development of molecular and serologic tests in a short period of time. While tests such as RT-PCR have applications in the immediate diagnosis revealing the presence of the virus, serological tests can be used to determine previous exposure to the virus and complement acute diagnosis. Antibody production can occur as early as 5 days post-infection. Both IgM and IgG specific anti-SARS-COV-2 antibodies can be a useful tool to test faster and larger groups of individuals. The objective of this study was to carry out a review of the different serological tests offered to detect antigen or antibodies against SARS-CoV-2. This information should be useful for decision takers in different countries to choose a test according to their needs. Based on web pages that listed serological assays, we found 226 coming from 20 countries, the majority are indirect tests for specific antibodies detection (n 180) and use immunochromatography methods (n 110) with samples coming from blood-derived products (n 105). Measuring IgM/IgG at the same time (n 112) and a procedure time of <20 min (n 83) are the most common. The overall average sensitivity was 91.8% and specificity was 97%. Most of the tests are currently for in vitro diagnosis (IVD). This information gathered could change day by day due to the expedite process of production and emergency of authorization use.

## Introduction

Nidoviruses are positive-sense single-stranded RNA viruses that infect a large number of vertebrates. Within these is the family of coronaviruses which has four groups, including betacoronavirus, that caused epidemic outbreaks in recent decades (1). Although coronaviruses were described as causing common respiratory symptoms in the 1960’s (2), they may be responsible for between 7 to 15% of uncomplicated upper respiratory infections (3). SARS (severe acute respiratory syndrome) in 2002 from China was the first report of a coronavirus outbreak with a mortality around 10%. The virus was probably transmitted to humans by a mammal (Civet cat) but probably originally derived in bats. The second outbreak was MERS (Middle East respiratory syndrome), originating in Saudi Arabia, transmitted by camels, but also originally derived in bats; with a mortality close to 40% (1). Now, we have a third epidemic, the coronavirus (CoV) SARS-CoV-2, which produces COVID-19 (coronavirus disease 2019) which began in the Wuhan province in China but has now turned into a pandemic. The sequence of the virus genome isolated from patients is similar to a bat virus (4). In China, the infection produced mild respiratory symptoms in about 80% of those infected, however, 5% were admitted to the Intensive Care Unit (ICU) of which 2.3% received mechanical ventilation and a mortality of 1.3% (5). The rapid case growth around the world means that in a short time the health systems with scarce resources, such as ICU teams, could saturate rapidly (6). The current standard assay for COVID-19 diagnosis is the detection of viral RNA using RT-PCR in nasopharyngeal swabs (7). Rapid and simple immunoassay tests have been developed to detect antigen or IgM and IgG antibodies (separately or simultaneously) against the SARS-CoV-2 virus in human blood even within 15 minutes. Antibody response can be detected as early as 5 days post-infection (8) and the antibody-secreting cells peak around day 7-8 post-infection (9, 10). One of the first peer reviewed studies of this kind of assays showed a test with a sensitivity of 88.66% and a specificity of 90.63% in 397 patients with SARS-CoV-2 confirmed by PCR (11). There are currently more than 200 immunoassays for SARS-CoV-2 to detect antigens or specific antibodies (12). The goal of this study is to carry out a comprehensive review of the wide offer of serological kits to detect SARS-CoV-2 antigen or antibodies, in order to help institutions and policymakers define the best option for a possible massive testing. There is an urgent need for rapid serological tests for SARS-CoV-2 that will be a useful tool for public health in the upcoming days.

## Methods

Two approaches were used for the literature search, web searches for pages listing serology tests for SARS-COv-2 and Pubmed (https://www.ncbi.nlm.nih.gov/pubmed/) for peer reviewed literature. Descriptive information from each test was obtained from technical data sheets (TDS) or in their respective company web page. Variables obtained were: country of origin, type of immuno-assay, procedure time, sample type, fixed antigen and antibodies isotype for indirect assays, sensibility, specificity, current regulatory status and published studies. We used not reported (N/R) to specify when information about a variable was not found; and N/A when a variable does not apply. A Pubmed search was conducted for articles describing studies of serology with human samples for SARS-CoV-2. Keywords used were: human + serology + either nCoV, SARS-CoV-2 and Covid-19 or human + antibodies + either nCoV, SARS-CoV-2. We examined the articles, looking for ones that mentioned the use of commercial antigen or antibody detection kits. Data was obtained until April 5th 2020.

### Data analysis and report

Information was stored in an Excel file (Microsoft, Redmond, WA). Data was randomly chosen to be verified by two authors. Information was presented as percentages, and means. No statistical analysis was applied.

## Results

We scanned the internet for web pages listing immunoassays for SARS-CoV-2, until april 5th of 2020, and four were used: https://www.finddx.org/ (n 213), https://www.modernhealthcare.com/ (n 99), https://www.fda.gov/ (n 54) and https://www.minsal.cl/ (n 12), **Supplementary Table 1**. The last web page was included because it had information about assays developed in South America, not found in the other lists. Information about kits and companies were crossed to complete or eliminate entries. We used companies web pages and technical data sheets (TDS), the official document provided by the manufacturer including specific characteristics, and instructions for use, clinical and analytical performance. In total, we found an offer of 226 immunoassays from 20 different countries, of these, 80.1% came from 3 countries, China, USA and South Korea. These countries represent 48.7%, 21.7% and 9.7% of the tests, respectively. TDS’s were found in 22.1% of tests. Serological tests were divided in antigen detection (direct) or antibody detection (indirect), according to each TDS. When TDS was not available, it was assigned according to the description of the test’s name. Only 3.1% were not found. From the reported, 82.2% were indirect and 17.8% were direct tests. Samples used to carry out the assays were categorized in: blood-derived (blood, serum, plasma) and naso-oropharyngeal swab and other fluids (oropharyngeal swab, bronchoalveolar lavage or sputum). For 112 of the tests, the sample type was not identified and from the reported, 92.1% use blood-derived samples, and only 7.9% swabbing samples. All of the samples obtained from naso-oropharyngeal swabs were for direct tests.

From the antigen assays (direct n 39), only two reported the specific antibodies fixed to the plate, antibodies against viral nucleocapsid (N) protein. From the antibody assays (indirect n 180) 25 reported the antigen use for antibody detection: 2 whole viral antigen and 23 recombinant proteins including the spike (S) protein (n 3), nucleocapsid (n 1) and recombinant unspecified viral antigen (n 19). From 172 assays reported the method used for evaluation. Most of these tests were based on immunochromatography (63.6%), followed by ELISA (23.1%) and different methods for fluorescence detection (6.4%). There are also techniques based on chemiluminescence, immunoturbidimetry and bioelectronic detection. Of the total group of antibody tests, some analyzed a unique antibody isotype: IgA one test (0.5%), IgG 22 tests (11.7%) and IgM 24 tests (12.8%); others analyzed two isotypes simultaneously: 112 tests detected IgM/IgG (59.6%), 2 IgM/IgA (1.1%) and 2 measured three isotypes, IgA/IgG/IgM (1.1%). A total of 5 tests (2.2%) reported measurement of total antibodies, **Table 1**. Procedure time was not found in 124 (55%) and 102 (45%) reported the specific time. Of these, 43 test are done in 10 min or less, 40 between 10 to 20 min (42.2% and 39.2%, respectively), 4 between 20 to 30 minutes (3.9%), one between 30 min and 1 hour (1%), and 14 take more than an hour (13.7%), some of them reaching even two hours. For the tests that describe the strategy of test interpretation (n 166), most of the assays (65%) are immunochromatography, results that are visualized as bands, and 35% are automated.

**Table 1.** Antibody isotypes measured by indirect serological assays.

| **Antibody isotype** | **n** | **Percentage (%)** |
| --- | --- | --- |
| Total antibodies | 5 | 2,7 |
| IgA | 1 | 0.5 |
| IgG | 22 | 11.7 |
| IgM | 24 | 12.8 |
| IgA/IgM | 2 | 1.1 |
| IgM/IgG | 112 | 59.6 |
| IgA/IgG/IgM | 2 | 1.1 |
| N/R | 20 | 10.6 |
| **Total** | **188** | **100** |

N/R: Not reported

A total of 18.6% of the assays reported internal validation defined here as the data either found in the TDS or provided by the manufacturer web pages. The number of assays that reported sensitivity or specificity was 41 (18,1%) and 42 (18,6%) respectively. The sensitivity of the tests ranged between 45% and 100%, **Table 2**. On the other hand, the specificity of the assays ranged between 90.3% and 100%, **Table 3**. Only 33 (14.1%) provided the number of donors evaluated for the previous analysis, the average number of donors tested was 274 and the average percentage of infected individuals on those was 36.1%. More specific data on sensitivity, specificity and the number of people tested can be found in **Supplementary Table**

**Table 2.** Reported sensitivity of the serological assays (internal validation)

|  | **IgM** | **IgG** | **IgG+IgM** |
| --- | --- | --- | --- |
| ≤ 80% | 6 (23.1%) | 2 (7.7%) | 1 (5.2%) |
| 80 - 90% | 15 (57.7%) | 6 (23.1%) | 3 (15.8%) |
| 90 - 95% | 3 (11.5%) | 4 (15.4%) | 9 (47.4%) |
| 95 - 97.5% | 1 (3.8%) | 4 (15.4%) | 3 (15.8%) |
| ≥ 97,5% | 1 (3.8%) | 10 (38.5%) | 3 (15.8%) |
| **Total** | **26 (100%)** | **26 (100%)** | **19 (100%)** |

**Table 3.** Reported specificity of the serological assays (internal validation)

|  | **IgM** | **IgG** | **IgG+IgM** |
| --- | --- | --- | --- |
| ≤ 90% | 0 (0%) | 0 (0%) | 0 (0%) |
| 90 - 95% | 1 (4%) | 1 (4.2%) | 3 (16.7%) |
| 95 - 97.5% | 7 (28%) | 4 (16.7%) | 7 (38.9%) |
| ≥ 97,5% | 17 (68%) | 19 (79.2%) | 8 (44.4%) |
| **Total** | **25 (100%)** | **24 (100%)** | **18 (100%)** |

According to the information provided regarding to the intent to use and regulatory status; no current label requirement was found for 46 tests (20.4%) and most of the tests 113 (50%) with reports qualified for in-vitro diagnostics (IVD), 37 (16.4%) for research use only (RUO), 29 (12.8%) were in development and 1 (0.4%) as IVD/RUO. Only 23 have received regulatory certification (of emergency use) issued by health authority of each country; Australia (TGA) 3, Brazil (ANVISA) 6, China (NMPA) 4, European Community (CE) 6, India (ICMR) 1, Singapore (HSA) 2 and USA (FDA) 1, and 56 notified to FDA.

Finally, we reviewed the literature published until april 5th 2020, where they used serological assays listed in the **Supplementary Table 2**. From 15 articles, 4 kits were used among those publications (9, 11,13,14).

## Discussion

Although the current standard for SARS-CoV-2 is the amplification of viral RNA by RT-PCR, this technique requires special equipment and trained individuals (7). Also, detection of the virus is dependent on the sample origin and time of sampling (15, 16). Detection of anti-virus specific SARS-CoV 2 antibodies could help to determine the exposure of a large population to the virus (5, 8-10). In infected individuals, antibody detection by ELISA using nucleocapsid protein as antigen was identified at day 5 for IgM and for IgG 14 days (17). IgM antibodies are known to be produced early during a viral infection, followed up by the presence of IgG which have a longer life-span and are responsible for the memory response (18). During the mitigation phase, besides the diagnosis of the virus, it is important to determine the immune status of the individual against the SARS-CoV-2 using detection of specific antibodies. Now there is an offer of more than 200 rapid diagnosis tests including detection of viral antigen or specific antibodies, all of them in different stages of development.

Initial reports of the new CoV causing acute respiratory distress syndrome came up in Wuhan, China. Since then, several assays have been developed in order to improve the diagnosis, most of them from China (4, 19). As expected most of the available tests detect antibodies using blood-derived samples. Although RT-PCR is considered the most sensitive detection method in respiratory fluid samples, it increases the risk of contamination of healthcare workers (20). Blood-derived samples are easier to obtain, and compared to RT-PCR, serological tests are faster, require less training and no equipment, so they can be used in almost any setting (11). The most common method behind is lateral flow immunochromatography (21) since these tests have a long shelf-life, do not require refrigeration and easily distinguishable visual results exclude the need for additional equipment compared with other methods like ELISA (22). Antigens used for detection are very important; the genome of SARS-CoV-2 codifies for several structural proteins, including the spike (S), membrane (M), envelope (E), and nucleocapsid (N) proteins (23). The most common antigens used for indirect assays are the recombinant spike and nucleocapsid proteins (16, 17). The S protein contains the domain for attachment to the human host cells (24), and the nucleocapsid protein is one of the major structural components involved in many processes of the virus including viral replication, transcription, and assembly (18). Interestingly, there is a 90.5% homology among nucleocapsid proteins of SARS-Cov-1 y SARS-CoV-2 (17), and SARS-CoV-2 showed an homology of about 85% with a coronavirus isolated from bats (4, 16). Most tests assessed levels of IgG and IgM simultaneously. The humoral response to some SARS-associated coronavirus shows a simultaneous increase in the level of all antibodies (25). Also, the dual detection of IgG-IgM improves the sensibility in comparison with individual IgG or IgM antibody assay (11), suggesting a possible improvement in the infection detection. In terms of efficiency, a shorter procedure time means a greater evaluation capacity of a population.

Test specificity and sensitivity are key for determining the role of these assays in diagnosis and public health programs (26). Unfortunately, only a minority of the tests present this information, maybe due to short time for development. However, most of them present a sensitivity and specificity well over 90%, but with a low number of infected individuals. Patients with RT-PCR confirmed virus have a median seroconversion rate of 93.1% for IgG and IgM (9) in a time dependent manner (9, 11), and 15 days after disease onset seroconversion was 100% for both antibody isotypes (9). Meanwhile, the detection rate of molecular based methods decreased to as low as 45% in that same time (9, 15, 27). The latter shows that the specificity and sensitivity of each immunoassay are variable depending on the time of onset, with more positive results given in a later time of disease onset (17). Additionally, the different samples that can be used for serological diagnosis offer more consistent results, no significant differences were found in blood, serum or plasma samples (11). This is opposed to the great variability from samples used in viral RNA detection (sputum, naso/oropharyngeal swabs and bronchoalveolar lavage) (15, 27, 28). Both RT-PCR with antibody assays have advantages, however the combination of both can provide more accuracy to the initial diagnosis of SARS-CoV-2 infection (11, 20). Moreover, current label requirements showed that most of the offered assays (50%) are intended for in-vitro diagnosis and this application represents the usefulness in a clinical environment. At this point of the pandemic, it would be difficult to suggest which tests are the best for clinical application. However, the information here presented sheds light into the large number of assays available, and the number increases day by day. This has to be used carefully, we suggest that researchers and policymakers focus on the ones with the most information available, such as a rigorous internal validation data, a well defined TDS, and are intended to be used for in-vitro diagnosis. More research is needed, especially studies that compare between different tests which provide more accurate information than studies with single assays (29). Yet, the initial data looks promising and immunoassays could help screen larger populations in less time, increasing the detection rate and increasing the testing capacity, one of the cornerstones needed to decrease the SARS-Cov2 spread. Recent studies show that convalescent patients have high levels of SARS-CoV2 neutralizing antibodies (NAbs), which increased with patient age (30). Interestingly, transfusion of convalescent plasma obtained from COVID-19 cases, improved clinical outcomes of patients with severe disease (31); thus suggesting that the antibodies produced by COVID-19 patients during the infection have a posterior protective effect. Large serological studies to detect virus-specific antibodies will be needed to determine the infected asymptomatic population and also could help to suspend social isolation in seropositive individuals.

## Data Availability

Data is avalaible in the supplementary table 2 and link to web pages

## Acknowledgments

The authors want to thank Manuel Franco MD, PhD from Pontificia Universidad Javeriana, Bogotá DC, Colombia for critical revision of the manuscript.

---

**Supplementary table 1.** Web pages used to search SARS-CoV-2 immunoassays.

| **Web page** | **Entity** | **Link** | **Number** |
| --- | --- | --- | --- |
| **FIND** | FIND: Foundation for Innovative New Diagnostics. A global non-profit organization driving innovation in the development and delivery of diagnostics. | https://www.finddx.org/covid-19/pipeline/?section=immunoassays#diag_tab | **213** |
| **Modern Healthcare** | An independent American publisher of national and regional healthcare news. | https://www.modernhealthcare.com/safety/coronavirus-test-tracker-commercially-available-covid-19-diagnostic-tests | **99** |
| **ISP-Chile** | Public Health Institute from the Health Ministry-Chile | https://www.minsal.cl/wp-content/uploads/2020/04/Lista-Test-Rapidos-Covid-al-03_04_2020.pdf | **12** |
| **FDA** | The Food and Drug Administration (FDA or USFDA). Federal agency of the United States | https://www.fda.gov/medical-devices/emergency-situations-medical-devices/faqs-diagnostic-testing-sars-cov-2 | **54** |

**Supplementary Table 2.**
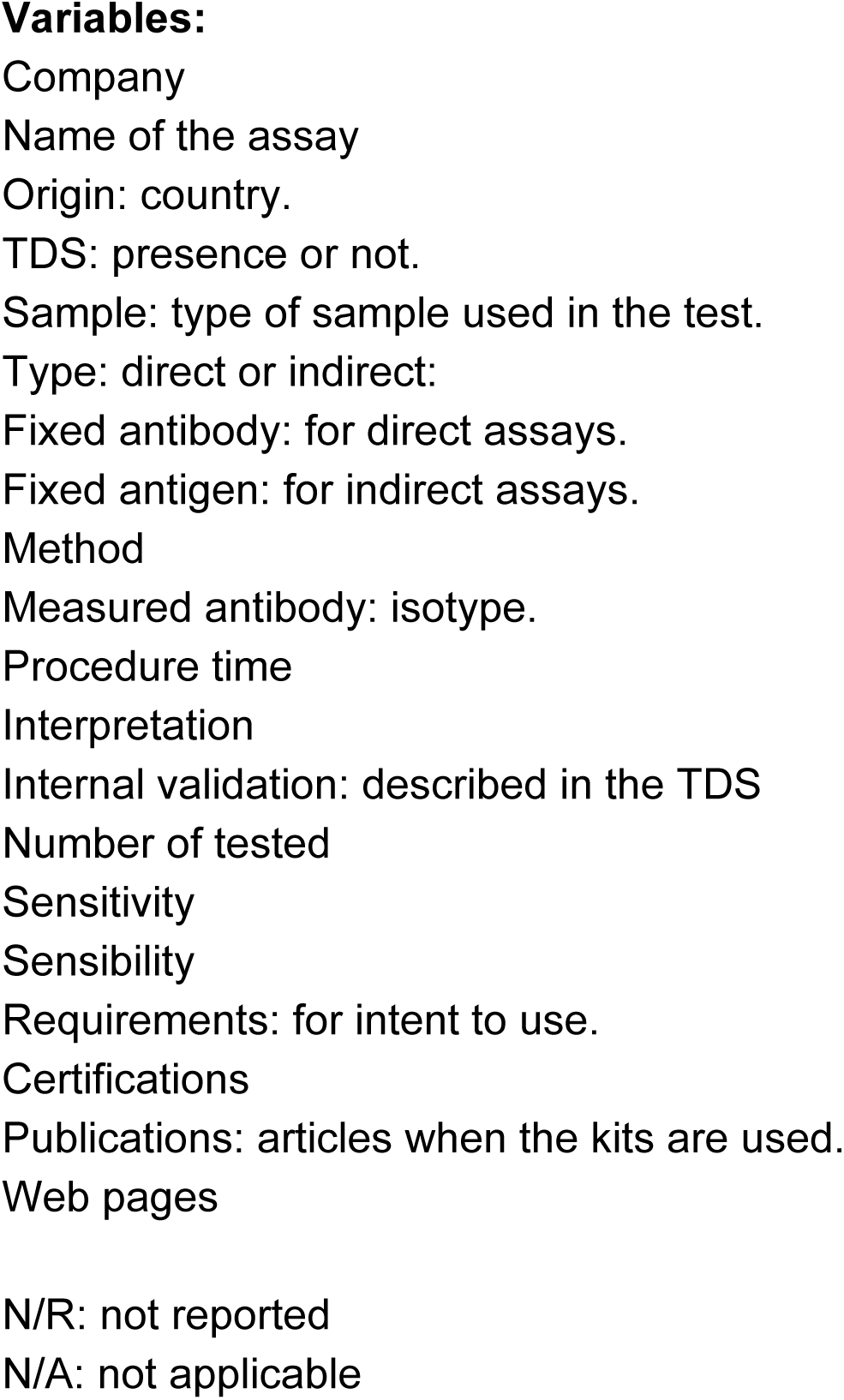
List of immunoassays analized (see annexed file).

